# A Logistic Curve in the SEIR Model and the Basic Reproduction Number of COVID-19 in Japan

**DOI:** 10.1101/2022.09.18.22279896

**Authors:** Takesi Saito, Kazuyasu Shigemoto

**Affiliations:** Department of Physics and Astronomy, School of Science, Kwansei Gakuin University, Sanda 669-1337, Japan; Tezukayama University, Nara 631-8501, Japan

**Keywords:** Variants of SARS-COV-2, SIR model, Epidemiology, Molecular biology

## Abstract

The SEIR model is one of modified models of SIR, especially taken into account of exposed people. SEIR equations can be solved numerically, but it is hard to obtain analytically. Here, we propose some approximate solutions of SEIR equations, one of which is related with the logistic formula in Biology. As the second aim, the SEIR model is applied to the 7th-wave of COVID-19 in Japan. The basic reproduction number (*α*) in the SEIR model is estimated for the Omicron wave. We make use of data of the removed number *R*(*t*) rather than that of the infective number, because the latter seems to be ambiguous. This analysis gives *α* = 10 with *γ* = 1 and σ = 0.5.

## 1. Introduction

The SIR model [1] in the theory of infection is powerful to analyze an epidemic about how it spreads and how it ends [3-16]. In this article we consider the SEIR model [2] which is one of modified models of SIR, especially taken into account of exposed people. SEIR equations can be solved numerically, but it is hard to obtain analytically. In order to analyze infections, analytic solutions are important. Here, therefore, we propose some approximate solutions of SEIR equations, one of which is related with the logistic formula in Biology.

As our second aim, the SEIR model is applied to the 7th-wave of Omicron-COVID-19 in Japan. The basic reproduction number (*α*) in the SEIR model is estimated for the 7th - wave. We make use of data of the removed number *R*(*t*) rather than that of the infective number, because the latter seems to be ambiguous. According to this analysis we have *α* = 10 with *γ* = 1 and *σ* = 0.5, where *γ* and *σ* are the removed and the exposed ratios, respectively.

## 2. A logistic formula from the SEIR model

The sequential SEIR equations are given by

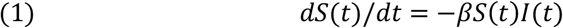

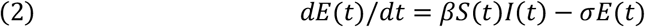

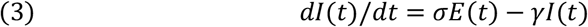

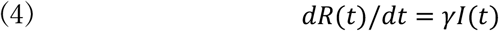

where *S*(*t*), *E*(*t*), *I*(*t*) and *R*(*t*) are numbers for susceptible, exposed, infectious and recovered, respectively, *α* = *β*/*γ* the basic reproduction number, *σ* the exposed ratio and *γ* the removed ratio. Four numbers are normalized as

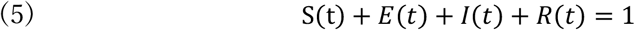

The SEIR equations can be solved numerically, when parameters *β, γ and σ* are given. In Fig.1 we give four curves for *S*(*t*), *E*(*t*), *I*(*t*) and *R*(*t*) with *β* = 10, *γ* = 1 *and σ* = 0.5.

Here we propose some approximate solutions of SEIR equations, one of which is related with a logistic formula in Biology. From Eqs. (1) and (4) we get *dS*/*dR* = −*αS*, which is integrated to be

**Fig. 1.**
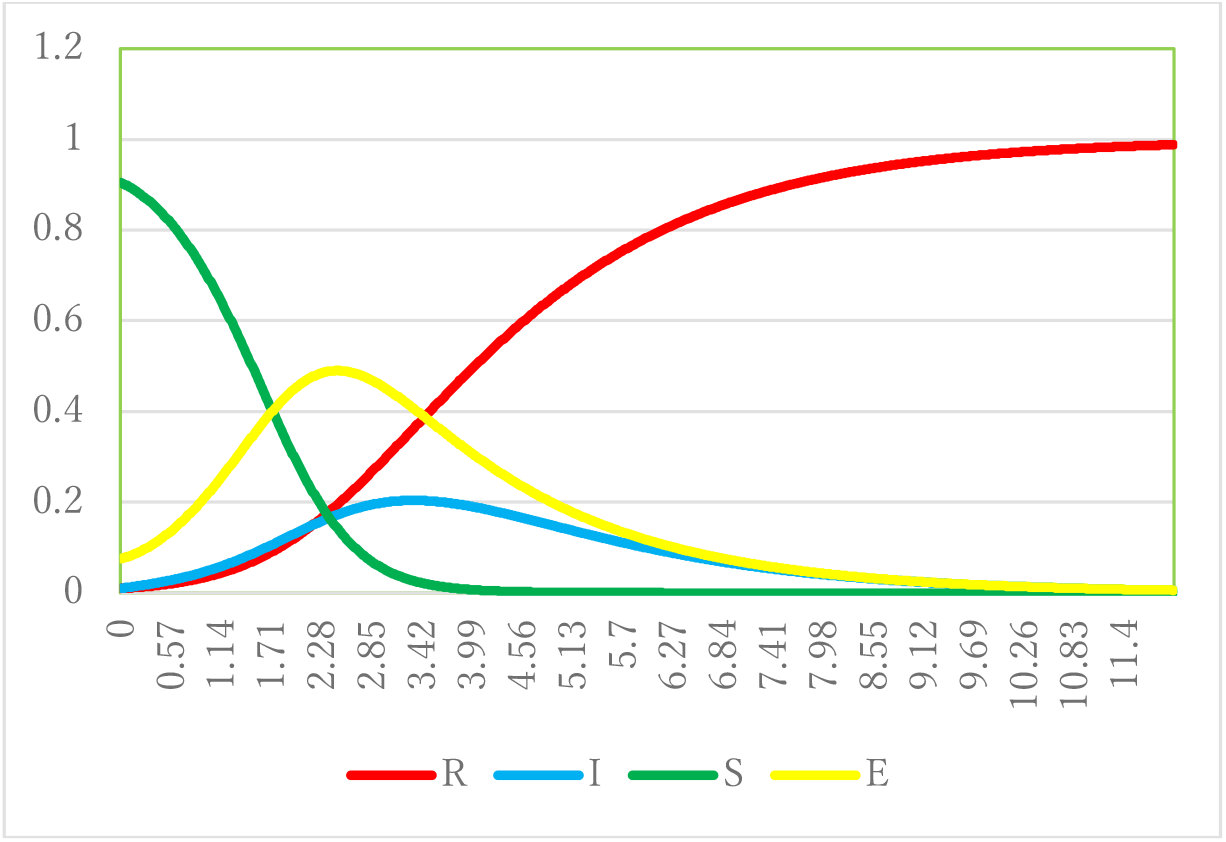
Four curves for *S*(*t*), *E*(*t*), *I*(*t*) and *R*(*t*) with *β* = 10, *γ* = 1 *and σ* = 0.5.

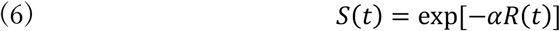

Hence we have from Eq. (5)

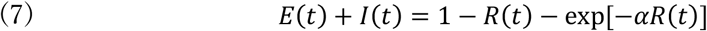

Now, from Eq. (3) *E*(*t*) can be expressed as

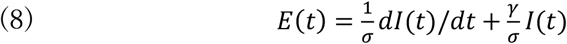

Substituting *I*(*t*) = *γ*^−1^*dR*(*t*)/*dt* of Eq. (4) into Eq. (8) above, we get

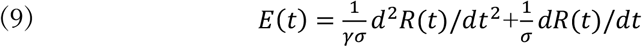

so that Eq. (7) reduces to the second order differential equation of *R*(*t*).

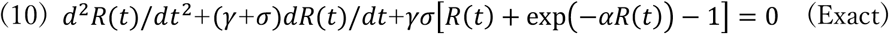

In order to find approximate solution of the exact equation (10), we consider three cases: Case 1: |*d*^2^*R*(*t*)/*dt*^2^ |≪ |*dR*(*t*)/*dt* |, *R*(*t*)

In this case the first term in Eq. (10) can be neglected so that we have

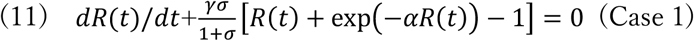

When *σ* ≫ *γ*, we find that this equation reduces to the SIR equation:

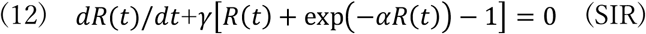

Case 2: When *αR*(*t*) ≪ 1, we can use the approximate formula exp(−*αR*) ≅ 1− *αR* + *α*^2^*R*^2^/2, then Eq. (10) reduces to

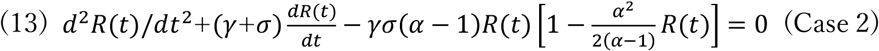

Case 3: A combination of Case 1 and Case 2 gives

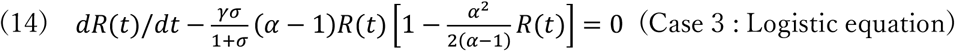

In Fig. 2, we draw curves of approximate solutions for Cases 1, 2 and 3, against the Exact equation (10). Here we find that the contribution of the second order differential part, *d*^2^*R*(*t*)/*dt*^2^, into any case is very small. The last equation (14), Case 3, is nothing but the logistic equation.

**Fig. 2.**
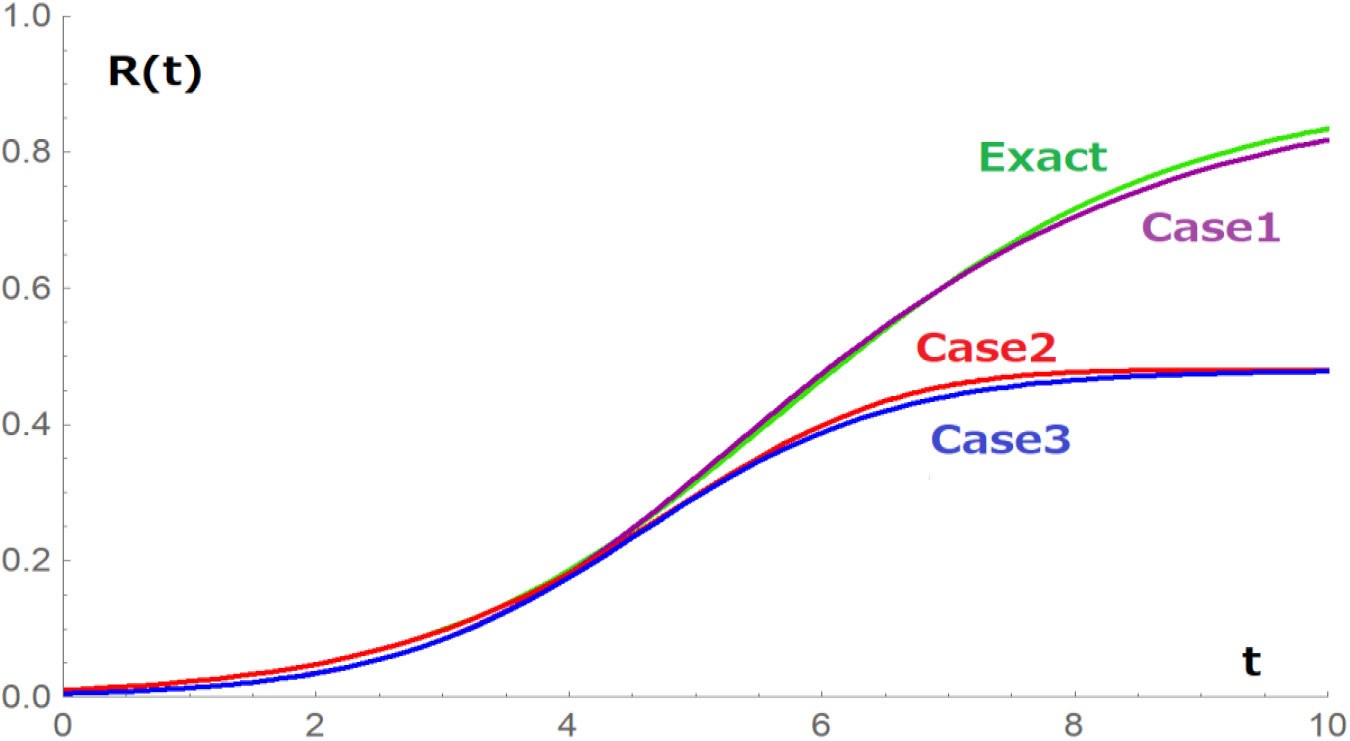
Curves of approximate solutions for Cases 1, 2 and 3, against the Exact equation. solution. The Case 3 is for the logistic equation.

## 3. The basic reproduction number of COVID-19 in Japan

Let us estimate the basic reproduction number in the SEIR model for the 7th-wave of COVI-19 in Japan. We make use of data [17] of the removed number *R*(*t*) rather than that of the infective number, because the latter seems to be ambiguous. In Fig. 3, the red curve of

**Fig. 3.**
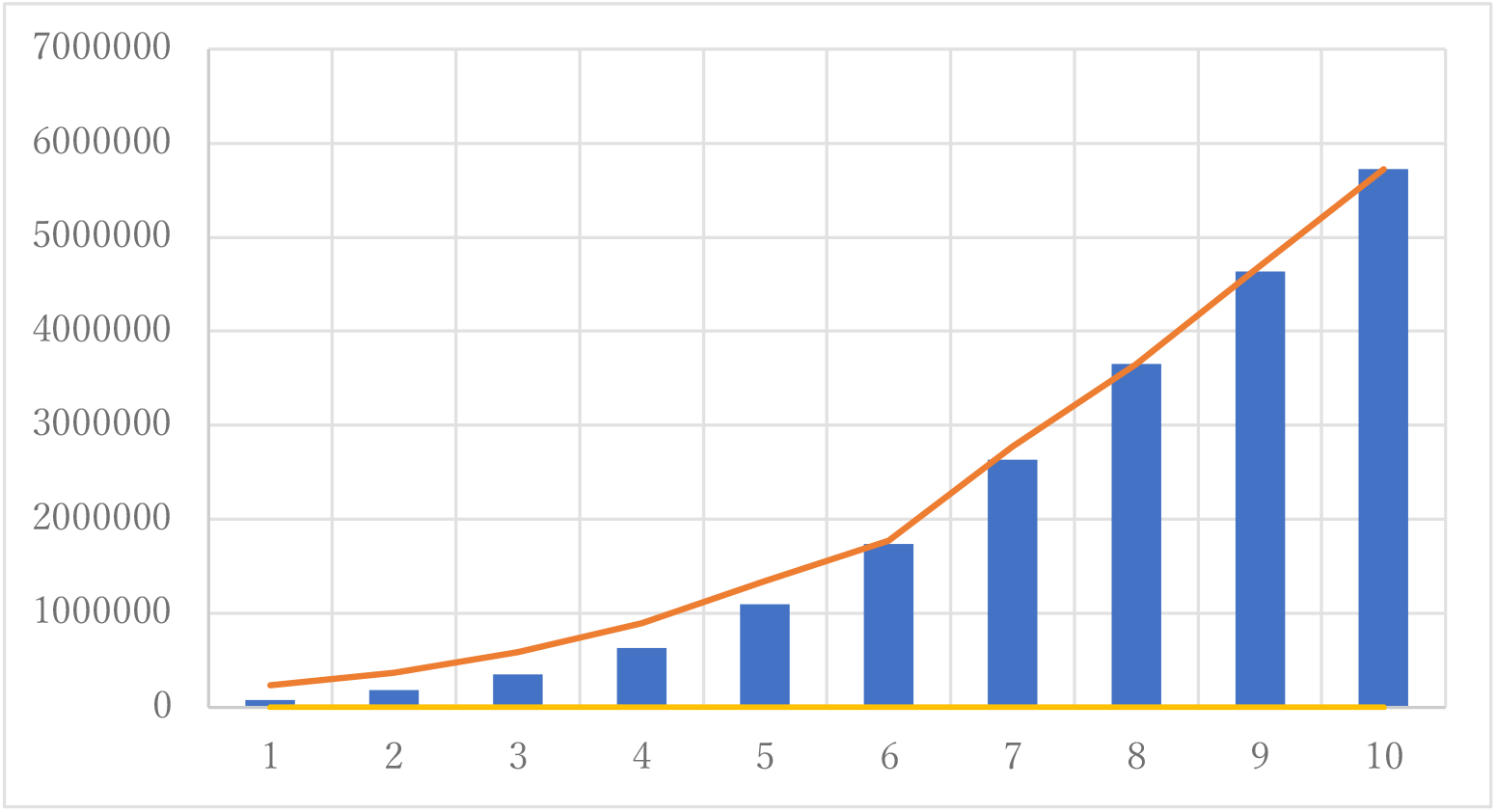
The red line shows a curve of *R*(*t*) with *α* = 10, *γ* = 1 *and σ* = 0.5. Blue bar charts stand for the 7th-wave data of *R*(*t*).

*R*(*t*) for the 7th-wave is drawn with *α* = 10, *γ* = 1 *and σ* = 0.5 in the SEIR model. Blue bar charts stand for the 7th-wave data of *R*(*t*). The number on the horizon axis corresponds with the date for each in Table 1.

**Table 1.** The number above corresponds with the date below for each.

|  |  |  |  |  |  |  |  |  |  |  |
| --- | --- | --- | --- | --- | --- | --- | --- | --- | --- | --- |
| 0 | 1 | 2 | 3 | 4 | 5 | 6 | 7 | 8 | 9 | 10 |
| Jul.1 | Jul.6 | Jul.11 | Jul.16 | Jul.21 | Jul.26 | Jul.31 | Aug.5 | Aug.10 | Aug.15 | Aug.20 |

In Fig. 3, we see that a coincidence between both curves is fairly well. So, we conclude that the basic reproduction number in the SEIR model is *α* = 10 *withγ* = 1 *and σ* = 0.5 for the 7th-wave of COVID-19 in Japan.

## 4. Conclusion

We have found some approximate solutions of SEIR equations. First, when *σ* ≫ *γ* in Case 1, the SEIR equation reduces to the SIR equation (12). Approximate solutions are given by Case1, Case 2 and Case 3, and their curves are drawn in Fig. 2, where Case 3 is known to be the logistic formula in Biology. Here we find that the contribution of the second order differential part, *d*^2^*R*(*t*)/*dt*^2^, into any case is very small.

The logistic equation (14) can be rewritten as

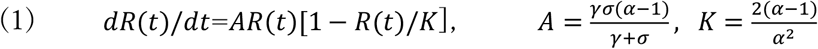

Then its explicit solution is easily obtained by

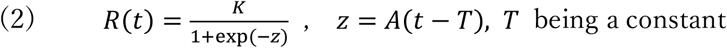

Hence, we have useful formulas

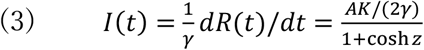

from Eq. (4) in Sec. 2, and

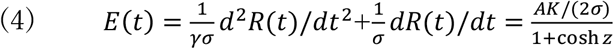

from Eq. (9) in Sec. 2, by neglecting the second order differential part, *d*^2^*R*(*t*)/*dt*^2^.

The second aim is the estimation of *α*, the basic reproduction number in the SEIR model for the 7th-wave of Omicron COVID-19 in Japan. We have made use of data of the removed number *R*(*t*) rather than that of the infective number, because the latter seems to be ambiguous. We have obtained *α* = 10 *with γ* = 1 *and σ* = 0.5.

## Data Availability

All data produced are available online at Yahoo Japan News page "Summary of the New Coronavirus Infection"(in Japanese), https://news.yahoo.co.jp/pages/article/20200207
and Toyokeizai net page "Status of the Domestic New Coronavirus Infection"(in Japanese),
https://toyokeizai.net/sp/visual/tko/covid19/

https://news.yahoo.co.jp/pages/article/20200207

https://toyokeizai.net/sp/visual/tko/covid19/

## Acknowledgment

We would like to express our deep gratitude to Kokado A. for valuable discussions.

